# Development of a Biology-Informed Chemical Mixture Index for Oxidative Stress and Mortality in NHANES 2005-2010: A Survey-Weighted Quantile G-Computation Approach

**DOI:** 10.64898/2026.06.30.26356938

**Authors:** Yanelli Rodríguez-Carmona, Kelly M. Bakulski, Erika Walker, Xin Wang, Wei Hao, Bhramar Mukherjee, Sung Kyun Park

## Abstract

**Background:** Current chemical mixture approaches are largely data-driven without considering shared biological mechanisms among mixture components, highlighting the need for biology-informed approaches.

**Objectives:** We constructed an integrated measure of a chemical mixture’s oxidative stress potential and assessed its association with mortality in the US population.

**Methods:** The sample comprised 4,574 adults (≥20 years) from National Health and Nutrition Examination Survey (NHANES) 2005-2010. To obtain robust estimates, we performed 1,000 repeated random 50:50 splits into training and testing sets. In each training set, we used survey-weighted quantile g-computation to model serum gamma-glutamyl transferase (GGT), an oxidative stress biomarker, as a function of a 30-chemical mixture (blood metals, urinary polycyclic aromatic hydrocarbons (PAHs), pesticides, phenols/parabens, and phthalates), adjusting for sociodemographic, behavioral, and dietary factors. We then applied the fitted model from each training set to the corresponding testing set to derive the environmental risk score for oxidative stress (ERS_OS_), defined by predicted GGT values. Associations of ERS_OS_ with all-cause, cardiovascular, and cancer mortality over 11 years of follow-up were estimated in the testing sets using survey-weighted Cox proportional hazards models and summarized across the 1,000 repeated splits.

**Results:** Chemicals with the largest positive weights in quantile g-computation included mono-(2-ethyl-5-hydroxyhexyl) phthalate, mono-2-ethyl-5-carboxypentyl phthalate, 2-hydroxyfluorene, methyl paraben, and benzophenone-3; chemicals with the largest negative weights included mono-(2-ethyl-5-oxohexyl) phthalate and PAH metabolites (1-hydroxynaphthalene, 3-hydroxyphenanthrene, and 3-hydroxyfluorene). The median correlation between observed and predicted GGT in the testing sets was 0.43 (2.5^th^, 97.5^th^ percentiles: 0.40-0.48). A one standard deviation increase in ERS_OS_ was associated with a median hazard ratio of 1.60 (2.5^th^, 97.5^th^ percentiles: 1.01-2.57) for cardiovascular mortality. No associations were found for all-cause mortality or cancer mortality.

**Discussion:** The proposed survey-weighted quantile g-computation approach may help estimate biology-informed chemical mixture effects in complex survey data, supporting the potential utility for population-generalizable environmental mixture research.

## Introduction

Exposure to environmental toxicants from pollution was estimated to be responsible for approximately nine million premature deaths worldwide in 2019.^1^ Oxidative stress, alongside subsequent inflammatory responses, is one of the main biological mechanisms by which environmental toxicants could cause chronic conditions, such as cancer, cardiovascular, respiratory, and neurological diseases.^2^ Although humans are exposed to complex mixtures of pollutants, many environmental health studies still examine *>one chemical at a time.* Similarly, most mixture studies have examined one chemical class at the time, and only a few have considered chemical mixtures from multiple classes.^3, 4^ *The use of single-chemical and single-chemical class approaches may* limit our understanding of how chemical mixtures influence oxidative stress and health.

The study of environmental mixtures presents significant statistical challenges due to the high-dimensional nature of the data, as well as the high correlations among chemicals.^5^ New statistical methods have been developed for environmental mixtures while some have been adapted from other fields to address the limitations of traditional methods.^6^ Furthermore, additional statistical complexities arise when utilizing data from national-level surveys in environmental mixtures research, like the National Health and Nutrition Examination Survey (NHANES), due to its complex sampling design. Many studies using NHANES data have failed to account for sampling weights, resulting in inaccurate estimates and potentially erroneous conclusions about the study population.^7^

Quantile g-computation (Qgcomp) is one of many statistical methods available to estimate the overall effect of a chemical mixture without having to first estimate the effects of individual exposures.^8^ Unlike other well stablished mixture methods, Qgcomp is not constrained by the directional homogeneity assumption, allowing variable weights to take positive or negative directions.^9^ Since the study of multi-class biology-based chemical mixtures is an underdeveloped area of research, Qgcomp’s ability to consider effects in any direction makes it a valuable method for advancing mixture research.

Currently, there is no universal definition for classifying chemicals, and groupings often rely on organ or biological system effects, shared health outcomes, molecular mechanisms, or structural characteristics.^10^ Oxidative stress is considered a common biological pathway underlying the toxicity of a wide range of environmental exposures, even across different chemical structure classes.^11^ Thus, evaluating the oxidative stress potential of a joint environmental exposure burden offers a promising approach for a shared pathway for linking the health effects of chemicals from distinct structural classes. This concept is motivated by toxic equivalent quantity (TEQ) framework,^12^ which is used to assess the toxicity of various combinations of dioxin and dioxin-like compounds. In TEQ, the relative potencies of individual compounds are determined by assigning toxic equivalent factors relative to 2,3,7,8-tetrachlorodibenzodioxin (TCDD), and these are aggregated into a cumulative risk index as the sum of toxicity-weighted masses or concentrations.^12^ TEQ serves as a good example of a biology-informed integration approach for assessing health effects of chemical mixtures; however, its calculation requires available toxicological data for each component of the mixture.^5^

Thus, we aimed to construct an environmental risk score (ERS) that integrates multi-class chemical biomarker data available in NHANES with a biomarker of oxidative stress, gamma-glutamyl transferase (GGT). To build the ERS for oxidative stress (ERS_OS_), we used survey-weighted Qgcomp accounting for NHANES complex survey design features including sampling weights and clusters, ensuring inference generalizable to the US adult population. Next, to evaluate the public health relevance of ERS_OS_, we examined its association with all-cause, CVD and cancer mortality outcomes in the same population.

## Materials and Methods

### Study population

NHANES is a series of cross-sectional surveys based on a complex, multistage probability design to sample the civilian, noninstitutionalized United States population.^13^ Every year since 1999, NHANES has collected information on health, nutritional status, and health behaviors from representative samples of approximately 5,000 individuals. NHANES participants provided informed consent to the National Center for Health Statistics. Between 1999–2010, NHANES data collection components were modified biannually based on emerging public health priorities.^13^ Many measures are collected within subsamples of each survey year, rather than across all participants. For example, most environmental exposure biomarkers in NHANES were collected for a one-third subset of the participants for a particular cycle (labeled as A, B, and C for convenience). NHANES cycles 2005-2006, 2007-2008, and 2009-2010 were selected for this analysis due to their greater overlap in continuously measured chemical classes and their subsample-specific availability.

### Chemical biomarker measures

Participants provided blood and urine samples in the mobile exam centers. Specimens were stored frozen until shipped to National Center for Environmental Health for analysis. Full details of chemical measurements are available in the NHANES website.^14^ Values below the limit of detection (LOD) were replaced by the LOD of the chemical divided by the square root of two. We identified five chemical classes that were measured in the same set of participants for mixture analysis across NHANES cycles 2005-2010: metals (measured in blood for all participants), and chlorophenol pesticides, polycyclic aromatic hydrocarbons (PAH), phenols and parabens, and phthalates (measured urine in subsamples B across these cycles). Detailed analytical and quality control procedures are available on the NHANES website.^14^ Biomarkers with a detection rate ≥ 50% were included, resulting in 30 chemicals across these five chemical classes (blood metals, urinary chlorophenol pesticides, PAH, phenols and parabens, and phthalates).

### Gamma glutamyl transferase measurements

Gamma glutamyl transferase is a liver enzyme involved in glutathione metabolism, and a thiol responsible for protecting cells against oxidative stress.^15^ Serum GGT is therefore regarded as a surrogate biomarker for oxidative stress,^16, 17^ and was selected to represent a common pathway between environmental chemicals and disease in humans.^2, 17^ Serum GGT was measured using an enzymatic rate method. For the 2005-2006 cycle and in 2007, GGT activity was measured using a Beckman Synchron LX20. In 2008, there was a change in methods and a DxC800 system was used for 2008 and the following 2009-2010 cycle.^18–21^ In the reaction, GGT catalyzes the transfer of a gamma-glutamyl group from gamma-glutamyl-p-nitroaniline (colorless substrate) to the acceptor, glycylglycine, with production of p-nitroaniline (colored product). The activity of GGT was measured by the change in absorbance at 410 nm over a fixed-time interval.

### Mortality outcome measures

NHANES data were linked to the public-use National Death Index (NDI), providing mortality data for adults ≥ 20 years through the last update on December 31, 2019. The underlying cause of death was defined using the International Classification of Diseases 10th Edition (ICD-10). All-cause mortality was defined as death from any cause. Cardiovascular disease (CVD) included heart disease (ICD-10 codes I00-I09, I11, I13, I20-I51), and cerebrovascular diseases (codes I60-I69). Cancer mortality causes were defined using ICD-10 codes C00-C97. Follow-up time was calculated from the baseline NHANES interview date to the date of death or December 31, 2019, for the participants without an event.

### Covariate measures

Demographic characteristics such as age, sex, race, and education were collected using questionnaires. In the 2005-2006 cycle, participants aged 85 years and older were top-coded as 85 to protect confidentiality. In 2007-2010, the top-coding threshold was lowered to 80 years. Sex was categorized as male or female. Education level was categorized into less than high school, high school or some college, and college or above. Self-reported race and ethnicity were categorized as Non-Hispanic (NH) White, NH Black, Mexican American (MA) and other Hispanics, and other race or ethnicity. Race and ethnicity was included as a covariate due to its role as a potential confounder, as it is associated with disparities in environmental exposures,^22^ and differential patterns of inflammatory biomarker levels and mortality risk have been observed across racial and ethnic groups.^23^ Participants were divided into three categories based on their smoking status: never smokers (smoked <100 lifetime cigarettes), current smokers (smoked ≥100 lifetime cigarettes and currently smoke some days or every day), and former smokers (smoked ≥100 lifetime cigarettes but do not smoke now). Body mass index (BMI) was calculated by dividing the body weight in kg by the square of the height in meters and rounding the resulting values to one decimal.

Serum cotinine was measured by an isotope dilution-high performance liquid chromatography/atmospheric pressure chemical ionization tandem mass spectrometry. The LOD for serum cotinine was 0.015 ng/mL, and values below LOD were imputed with LOD/√2. Urinary creatinine concentrations were used to account for urinary dilution.

Dietary variables were considered as potential confounders because diet is associated with both lower and higher levels of oxidative stress biomarkers,^24^ and links have been suggested with lifespan and mortality outcomes.^25^ Additionally, diet represents an important pathway of exposure for several environmental chemicals.^26^ Two 24-hour dietary recalls were conducted by trained personnel at the NHANES mobile examination center.^27–29^ Observations with unreliable dietary data based on quality or completeness were dropped. Daily nutrient and total energy intakes (TEI) were averaged if two measurements were available. The Healthy Eating Index (HEI) 2015, a measure of the overall diet quality where a higher score (range 0-100) represents a higher adherence to the dietary guidelines,^30, 31^ was calculated ^32, 33^. Alcohol (g/day) intake from the diet recall was selected as a covariate because it contained fewer missing observations when compared to the alcohol intake frequency variables obtained from questionnaires (3.83% vs ∼20%, respectively).

### Statistical analysis

All analyses were performed in R statistical software (version 4.5.1),^34^ and code was reviewed by an independent analyst. All analyses were survey-weighted using the exposure subsample-specific weights and indicators of stratum and primary sampling unit (PSU) to retain national representativeness, unless otherwise specified. From the 31,034 participants from NHANES cycles 2005-2010, we included only participants part of subsample B (n=8,171). We further excluded those younger than 20 years of age (n=2,762) and removed observations with missing chemical biomarker measures (n=780), GGT measurements (n=49), and mortality data (n=6), for an analytic sample size of n=4,574 (**Figure 1**). To avoid introducing analytical complexity to the analysis, single imputation was performed using the mice package (version 3.19.0)^35^ for BMI (0.85% of unweighted missingness), cotinine (0.04% of unweighted missingness), smoking (0.07% of unweighted missingness), education (0.13% of unweighted missingness), and dietary variables (average energy intake, average daily alcohol intake and HEI 2015) (3.83% of unweighted missingness).

**Figure 1:**
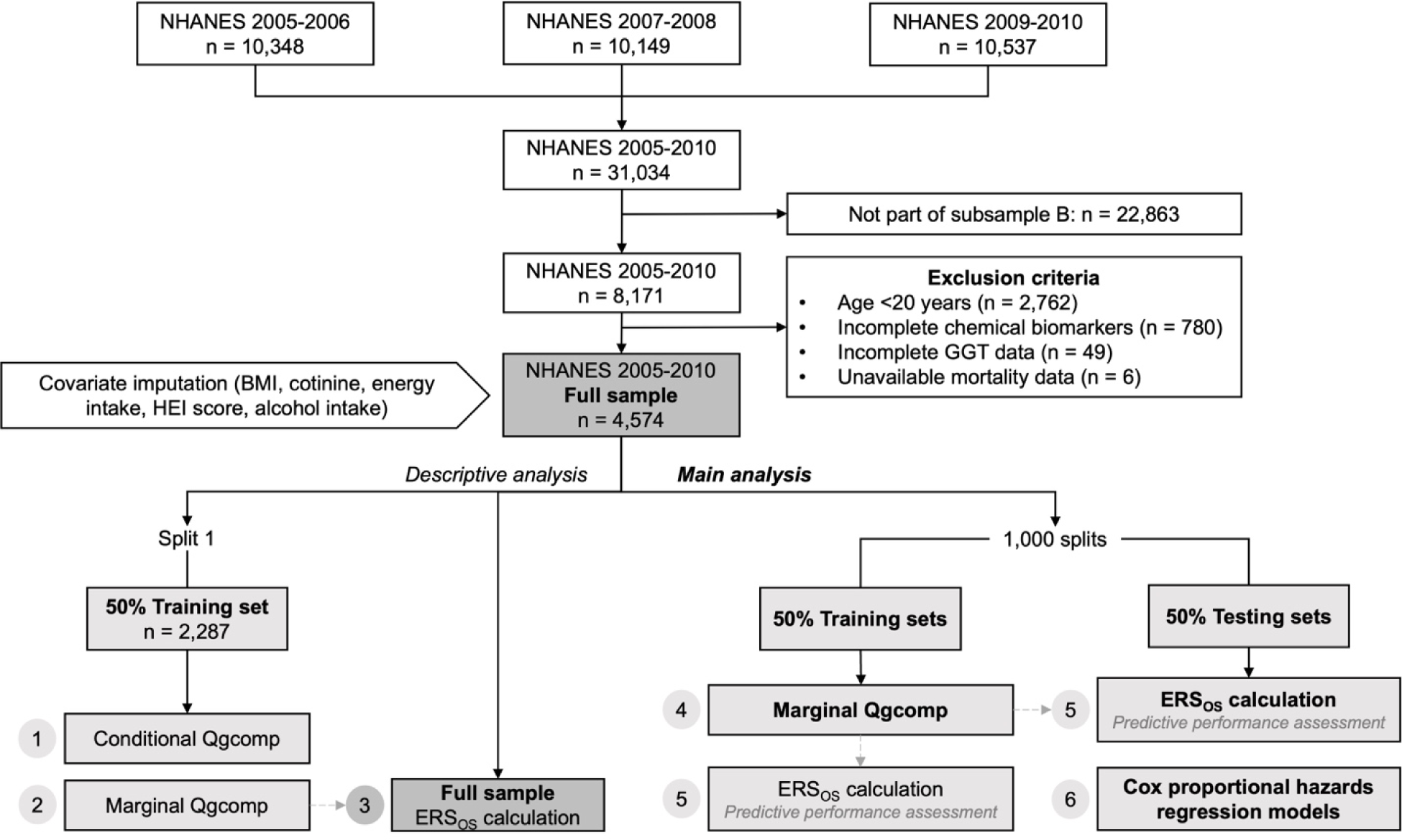
Sample size flowchart. Note: Dashed gray arrows depict the prediction of GGT values (ERS_OS_ computation) based on marginal Qgcomp models fitted in the training sets Abbreviation: BMI, body mass index; ERS_OS_, environmental risk score for oxidative stress; HEI, Healthy Eating Index; GGT, gamma-glutamyl transferase; NHANES, National Health and Nutrition Examination Survey; Qgcomp, quantile g-computation

We compared the distributions of participant characteristics between the included and excluded participants. For continuous variables, we used median, first quartile, and third quartile. For categorical variables, we used the unweighted number of participants and the survey weighted frequency, calculated with the survey package (version 4.4-8).^36^ GGT and cotinine were natural-log transformed. Concentrations of the 30 chemical biomarkers were log-transformed and Z-score standardized. In the full sample, we calculated unweighted pairwise Spearman correlations among the 30 chemical biomarkers included in the mixture and visualized the relationships using a heatmap in the full sample (n=4,574).

A single 50:50 training and testing data split was performed for visualization and descriptive analyses purposes. For the main analyses, to obtain robust estimates, we performed 1,000 repeated random 50:50 splits into training and testing sets (**Figure 1**). Briefly, in each training and testing set, we constructed ERS_OS_ using survey-weighted Qgcomp and evaluated the prediction performance of ERS_OS_. Finally, in each testing set, we assessed the associations between ERS_OS_ and mortality outcomes. The estimates obtained from the 1,000 splits are presented as medians and empirical 2.5^th^ and 97.5^th^ percentiles). Percentile ranges reflect split-to-split variability and should not be interpreted as formal 95% confidence intervals.

### Quantile g-computation

Qgcomp is a method to estimate the joint effect of a mixture on an outcome, rather than estimating each component’s contribution individually.^9^ Qgcomp first quantizes the exposures and then fits a generalized linear model to estimate the overall effect when all chemicals in the mixture are fixed at a certain quantile (i.e., mixture concentration increase by one quantile).^9^ During this procedure, fixed weights are calculated (Qgcomp weights). If all mixture chemicals have effects on the same direction, the obtained Qgcomp weights represent the proportion of the overall effect. However, if the mixture has chemicals in both positive and negative directions, each chemical individual Qgcomp weight represents the proportion of the mixture effect in a particular direction due to a specific exposure. The estimated Qgcomp weights in the same direction add up to one.^9^

In the single data split training set (n=2,287) (**Figure 1**), we performed a conditional model using the Qgcomp.noboot function (i.e. subject specific model) to visualize the direction of each of the chemical biomarker’s Qgcomp weights in the oxidative stress joint effect, as measured by GGT. The conditional Qgcomp was plotted using butterfly plots where the left bars represent the chemicals with negative effects on the outcome, and to the right are those with positive effects. Bar lengths represent the magnitude of each Qgcomp weight relative to the mixture effect size in the same direction. Chemicals with weights in the same direction to the overall effect size are presented in darker color shades. The data for the conditional Qgcomp was extracted and plotted using ggplot2 (version 4.0.1),^37^ where each of the 30 chemicals were color coded by chemical class. Furthermore, we also performed single pollutant survey-weighted regressions to evaluate the individual associations of the mixture components with GGT (n=2,287).

### Environmental risk score for oxidative stress (ERS_OS_) construction

To estimate the joint effect of the chemical mixture on oxidative stress, marginal Qgcomp models were fit in the 1,000 training datasets with GGT as the outcome, using the qgcomp.boot function from the Qgcomp package (version 2.18.7).^9^ The qgcomp.boot function estimates cluster-appropriate standard errors using nonparametric bootstrapping, which are then used to estimate Wald-type confidence intervals.^9^ The Qgcomp models were adjusted for age, sex, race/ethnicity, smoking, BMI, education, creatinine, cotinine, alcohol intake, energy intake, and HEI. We also accounted for sampling design (i.e. survey weights, strata and primary sampling units) by including the subsample sampling weights (subsample B) combined for NHANES 2005-2010 cycles (one third of wtsb2yr because three NHANES cycles were combined) and the product of stratum and the primary sampling unit (sdmvstra*sdmvpsu) as clusters. The resulting coefficients of the marginal Qgcomp (psi) represent the average change in log-transformed GGT concentrations, the oxidative stress measure, per one quartile increase in all chemicals simultaneously. The median and empirical 2.5^th^ and 97.5^th^ percentiles of psi across 1,000 repeated splits were back transformed to report the percent difference in GGT concentrations.

We computed the ERS_OS_ in the testing and training sets across 1,000 repeated splits by obtaining GGT predicted values from the marginal Qgcomp fitted in the training sets (**Figure 1**). Specifically, we used the internal msm.predict function to obtain predicted GGT values in the testing sets based on the qgcompfit objects generated when the qgcomp.boot function was called (marginal Qgcomp in the training sets).^9^ The resulting predicted values of GGT, the oxidative stress measure, constitute the ERS_OS_. As Qgcomp uses both negative and positive weights, a high-risk score indicates that individuals with high concentrations of positive coefficient chemicals and low concentrations of negative coefficient chemicals have a higher risk for oxidative stress.

A marginal Qgcomp was also fit in the single training set (n=2,287), and ERS_OS_ values were predicted for the full sample size (n=4,574) (**Figure 1**). Population characteristics were then compared by ERS_OS_ quartiles using *p*-trend for continuous variables and Pearson’s χ^2^ Rao & Scott adjustment for categorical variables.

### Assessment of predictive performance of ERS_OS_

To evaluate the predictive performance of ERS_OS_ across the 1,000 repeated splits, we first computed correlation coefficients between observed log-transformed GGT and predicted GGT values (ERS_OS_), in the training and testing sets. We then calculated mean squared error (MSE) in the training datasets and mean squared prediction error (MSPE) in the testing sets.

#### Associations between ERS_OS_ and mortality

The ERS_OS_ variables in the testing sets were z-score standardized, resulting in a mean of zero and a standard deviation of one. Furthermore, the ERS_OS_ variables were also categorized into survey-weighted quartiles. To evaluate the associations of ERS_OS_ with all-cause, CVD, and cancer mortality risk, we fit separate survey-weighted Cox proportional hazards regression models in the testing sets. Hazard ratios (HRs) were estimated per one-SD increase in ERS_OS_ when modeled continuously and by ERS_OS_ quartiles, using the lowest quartile as the reference. Models were adjusted for age, sex, race/ethnicity, smoking, BMI, education, creatinine, cotinine, HEI, TEI, and alcohol intake. Median HRs and empirical 2.5^th^ and 97.5^th^ percentiles across the 1,000 repeated splits are presented.

#### Sensitivity analysis

To evaluate the impact of covariate imputation we performed a complete case analysis (n=4,361) across 1,000 repeated splits where we calculated the ERS_OS_ and performed Cox proportional hazards regression models between ERS_OS_ and mortality outcomes.

## Results

In the full sample, the median age was 46.0 years old (Q1, Q3: 33.0, 59.0), and the most frequent race or ethnicity was Non-Hispanic White (70.4%), followed by Mexican American and other Hispanics (12.9%), and Non-Hispanic Black (10.7%). Around half of the population was female (51.4%), had never smoked (52.9%), and had completed high school or partial college education (54.5%) (**Table 1**). Chemical biomarker concentrations are presented in **Table 2**. Overall, the included participants (n=4,574) had higher GGT concentrations, were older, and were more likely to be Non-Hispanic White, and had higher BMI, higher average daily energy intakes, higher HEI, and higher alcohol consumption compared with excluded participants. Furthermore, all-cause and cancer mortality cases were less frequent in the final analytical sample (**Table S1**).

**Table 1.** Population characteristics in the National Health and Nutrition Examination Survey (NHANES) cycles 2005-2010 (n=4,574).

|  | **Median (Q1, Q3);**<br>**Unweighted N (weighted %)** |
| --- | --- |
| **Age (years)** | 46.0 (33.0, 59.0) |
| **Sex** |  |
| Male | 2,257 (48.6) |
| Female | 2,317 (51.4) |
| **Race/Ethnicity** |  |
| NH White | 2,253 (70.4) |
| MA/Other Hispanic | 1,246 (12.9) |
| NH Black | 870 (10.7) |
| Other | 205 (6.0) |
| **Education** |  |
| < High school | 1,291 (18.5) |
| High school/some college | 2,352 (54.5) |
| ≥ College | 931 (26.9) |
| **BMI (kg/m<sup>2</sup>)** | 27.5 (24.0, 32.0) |
| **Average TEI (Kcal/day)** | 1,991.0 (1,520.0, 2,566.5) |
| **HEI 2015 score** | 52.9 (43.6, 62.0) |
| **Cotinine (ng/mL)** | 0.1 (0.0, 29.0) |
| **Average alcohol intake (g/day)** | 0.0 (0.0, 9.4) |
| **Smoking status** |  |
| Never smoker | 2,424 (52.9) |
| Former smoker | 1,026 (22.3) |
| Current smoker | 1,124 (24.9) |
| **Urinary creatinine (mg/dL)** | 111.0 (63.0, 165.0) |
| **Mortality** |  |
| All-cause | 695 (11.0) |
| CVD | 217 (3.6) |
| Cancer | 154 (2.5) |
| **Follow-up (years)** | 11.3 (10.1, 13.1) |

Abbreviations: BMI, body mass index; CVD, cardiovascular disease; GGT, gamma glutamyl transferase; HEI, Healthy Eating Index; MA, Mexican American; NH, Non-Hispanic; TEI, total energy intake.

**Table 2.**
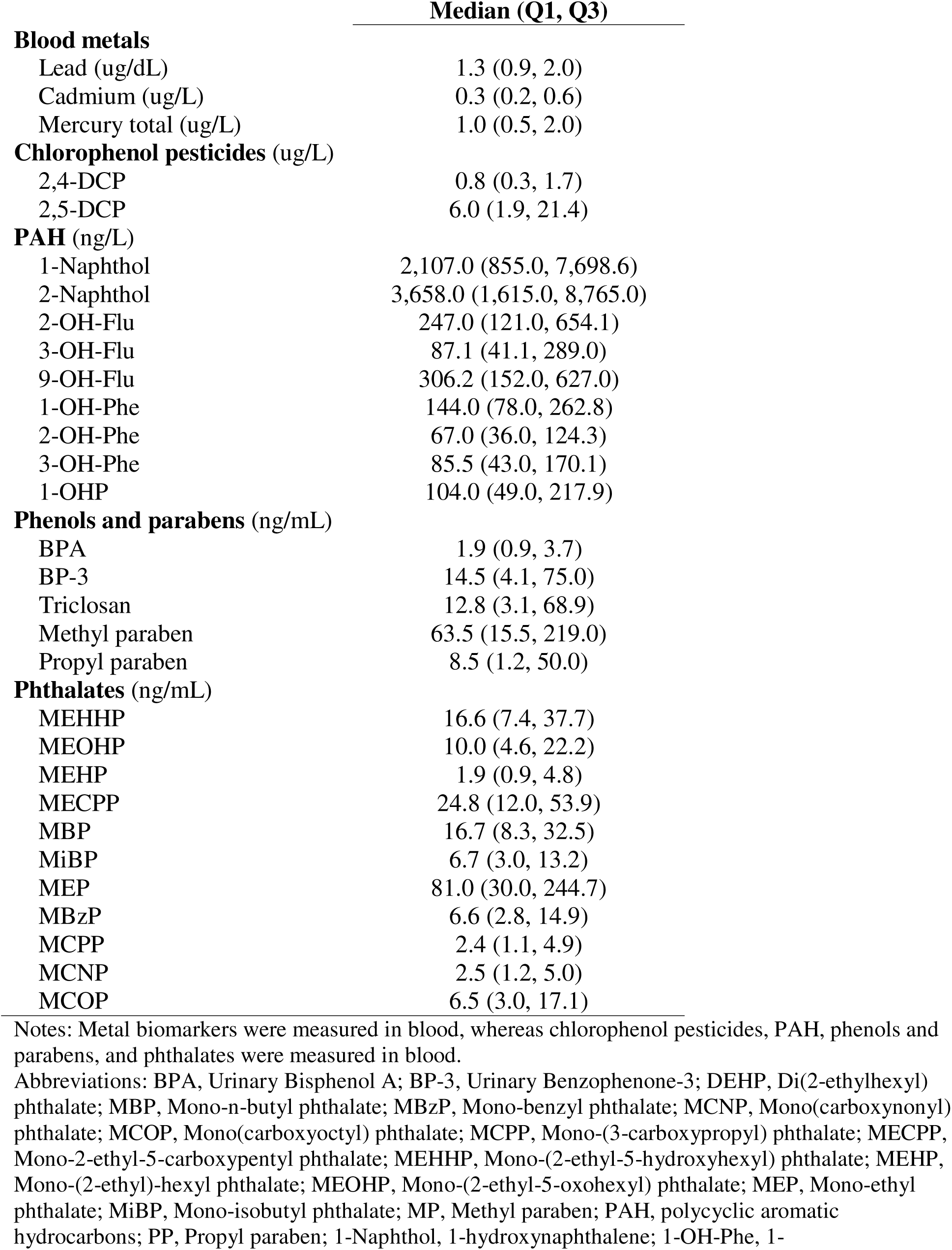
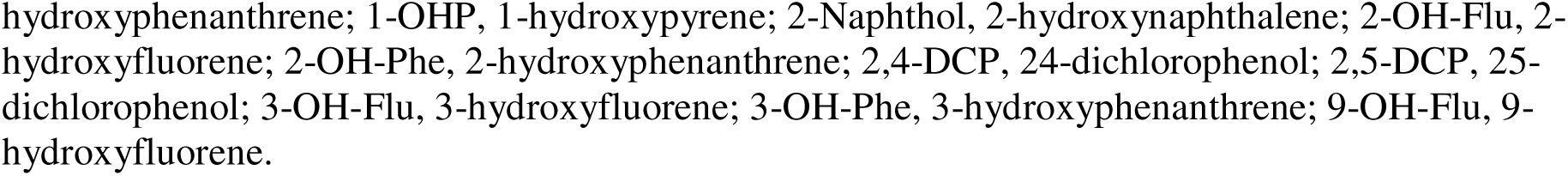
Distributions of chemical biomarker concentrations in the National Health and Nutrition Examination Survey (NHANES) cycles 2005-2010 (n=4,574).

|  | **Median (Q1, Q3)** |
| --- | --- |
| **Blood metals** |  |
| Lead (ug/dL) | 1.3 (0.9, 2.0) |
| Cadmium (ug/L) | 0.3 (0.2, 0.6) |
| Mercury total (ug/L) | 1.0 (0.5, 2.0) |
| **Chlorophenol pesticides (ug/L)** |  |
| 2,4-DCP | 0.8 (0.3, 1.7) |
| 2,5-DCP | 6.0 (1.9, 21.4) |
| **PAH (ng/L)** |  |
| 1-Naphthol | 2,107.0 (855.0, 7,698.6) |
| 2-Naphthol | 3,658.0 (1,615.0, 8,765.0) |
| 2-OH-Flu | 247.0 (121.0, 654.1) |
| 3-OH-Flu | 87.1 (41.1, 289.0) |
| 9-OH-Flu | 306.2 (152.0, 627.0) |
| 1-OH-Phe | 144.0 (78.0, 262.8) |
| 2-OH-Phe | 67.0 (36.0, 124.3) |
| 3-OH-Phe | 85.5 (43.0, 170.1) |
| 1-OHP | 104.0 (49.0, 217.9) |
| **Phenols and parabens (ng/mL)** |  |
| BPA | 1.9 (0.9, 3.7) |
| BP-3 | 14.5 (4.1, 75.0) |
| Triclosan | 12.8 (3.1, 68.9) |
| Methyl paraben | 63.5 (15.5, 219.0) |
| Propyl paraben | 8.5 (1.2, 50.0) |
| **Phthalates (ng/mL)** |  |
| MEHHP | 16.6 (7.4, 37.7) |
| MEOHP | 10.0 (4.6, 22.2) |
| MEHP | 1.9 (0.9, 4.8) |
| MECPP | 24.8 (12.0, 53.9) |
| MBP | 16.7 (8.3, 32.5) |
| MiBP | 6.7 (3.0, 13.2) |
| MEP | 81.0 (30.0, 244.7) |
| MBzP | 6.6 (2.8, 14.9) |
| MCPP | 2.4 (1.1, 4.9) |
| MCNP | 2.5 (1.2, 5.0) |
| MCOP | 6.5 (3.0, 17.1) |

Notes: Metal biomarkers were measured in blood, whereas chlorophenol pesticides, PAH, phenols and parabens, and phthalates were measured in blood.
Abbreviations: BPA, Urinary Bisphenol A; BP-3, Urinary Benzophenone-3; DEHP, Di(2-ethylhexyl) phthalate; MBP, Mono-n-butyl phthalate; MBzP, Mono-benzyl phthalate; MCNP, Mono(carboxynonyl) phthalate; MCOP, Mono(carboxyoctyl) phthalate; MCPP, Mono-(3-carboxypropyl) phthalate; MECPP, Mono-2-ethyl-5-carboxypentyl phthalate; MEHHP, Mono-(2-ethyl-5-hydroxyhexyl) phthalate; MEHP, Mono-(2-ethyl)-hexyl phthalate; MEOHP, Mono-(2-ethyl-5-oxohexyl) phthalate; MEP, Mono-ethyl phthalate; MiBP, Mono-isobutyl phthalate; MP, Methyl paraben; PAH, polycyclic aromatic hydrocarbons; PP, Propyl paraben; 1-Naphthol, 1-hydroxynaphthalene; 1-OH-Phe, 1-
hydroxyphenanthrene; 1-OHP, 1-hydroxypyrene; 2-Naphthol, 2-hydroxynaphthalene; 2-OH-Flu, 2-hydroxyfluorene; 2-OH-Phe, 2-hydroxyphenanthrene; 2,4-DCP, 2,4-dichlorophenol; 2,5-DCP, 2,5-dichlorophenol; 3-OH-Flu, 3-hydroxyfluorene; 3-OH-Phe, 3-hydroxyphenanthrene; 9-OH-Flu, 9- hydroxyfluorene.

Over a median follow-up time of 11.3 (Q1, Q3: 10.1, 13.1) years, 695 all-cause mortality cases, 217 CVD mortality cases, and 154 cancer mortality cases occurred (**Table 1**). Mortality cases recorded during the follow-up were on average older, were more likely to be non-Hispanic White, have less than high school education, lower total energy and alcohol intakes, and to be current smokers (**Table S2**). Furthermore, mortality cases had overall higher blood lead and blood cadmium concentrations, and slightly lower total mercury concentrations (**Table S3**). Most of PAH concentrations tended to be lower for all-cause and cancer mortality cases, except for 1-Napthtol, which was higher. Similarly, mortality cases also had lower concentrations for most phenols and parabens, and overall lower phthalate levels (**Table S3**).

Correlations within chemical classes in the mixture were mostly positive (**Figure S1)**. Correlations among blood metals ranged from very weak to weak (*r*=-0.07 for cadmium and total mercury, *r*=0.09 for lead and total mercury, and *r*=0.32 between lead and cadmium). A positive strong correlation was observed among chlorophenol pesticides (*r*=0.79), moderate to very strong positive correlations for PAH (*r*=0.53 to 0.96), very weak to very strong positive correlations among phenols and parabens (*r*= 0.15 to 0.82), and very weak to very strong positive correlations among phthalates (*r*=0.17 to 0.98) (**Figure S1**).

### Quantile G-computation and ERS_OS_ calculation

Weights obtained with Qgcomp represent the proportion of positive or negative partial effects of each chemical biomarker on the oxidative stress outcome, GGT. In the conditional Qgcomp model performed in the single testing set (n=2,287), the environmental chemicals with the largest positive weights included phthalates such as Mono-(2-ethyl-5-hydroxyhexyl) phthalate (MEHHP) and Mono-2-ethyl-5-carboxypentyl phthalate (MECPP), a PAH metabolite, 2-hydroxyfluorene (2-OH-Phe), and two parabens, methyl paraben (MP) and Benzophenone-3 (BP-3) (**Figure 2**). On the other hand, chemicals with the largest negative weights included one phthalate, Mono-(2-ethyl-5-oxohexyl) phthalate (MEOHP), followed by PAH metabolites like 1-hydroxynaphthalene (1-Naphtol), 3-hydroxyphenanthrene (3-OH-Phe), and 3-hydroxyfluorene (3-OH-Flu) (**Figure 2**). Single-pollutant associations between the biomarkers in the chemical mixture and GGT are presented in **Table S4**. Positive associations were observed between GGT, blood lead, and three phthalates, MEHHP, MECPP, and Mono-isobutyl phthalate (MiBP), of which two are Di(2-ethylhexyl) phthalate (DEHP) metabolites (MEHHP, and MECPP). Conversely, only 9-OH-Flu, a urinary PAH metabolite, was inversely associated with GGT (**Table S4**).

**Figure 2:**
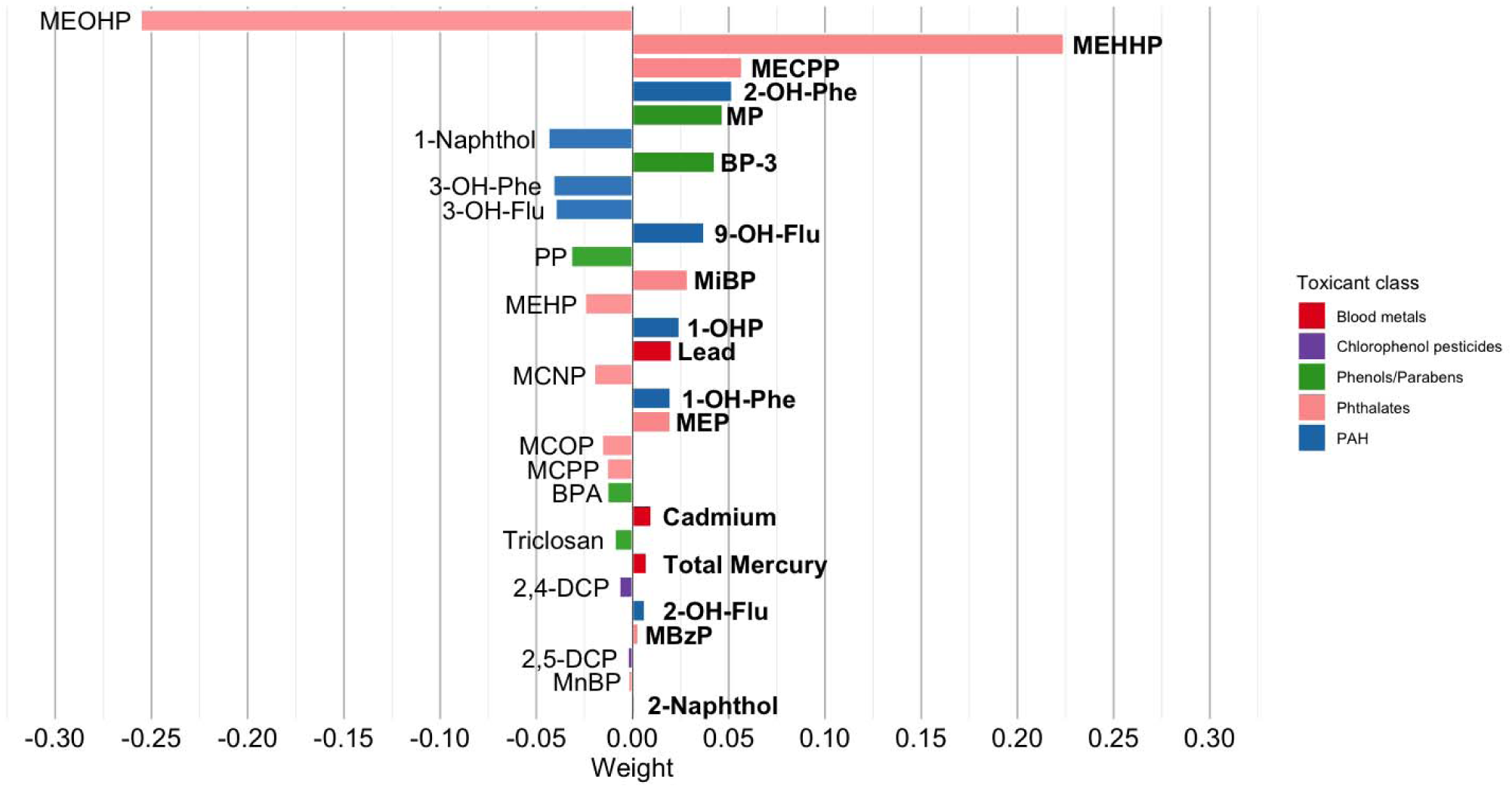
Conditional quantile g-computation model for oxidative stress in the training set (n=2,287). Notes: Quantile g-computation weights represent the proportion of positive or negative partial effects of each chemical on gamma-glutamyl transferase (GGT), the oxidative stress biomarker. Thus, bar size indicates relative weights within each direction. Metals biomarkers were measured in blood, whereas chlorophenol pesticides, PAH, phenols and parabens, and phthalates were measured in blood. Qgcomp weights in the positive direction had the greatest coefficient sum (sum of positive coefficients = 0.591, sum of negative coefficients = −0.51). The overall effect size was positive at 0.081. The bold font reflects the overall positive “mixture” effect. Abbreviations: BPA, Urinary Bisphenol A; BP-3, Urinary Benzophenone-3; DEHP, Di(2-ethylhexyl) phthalate; MBP, Mono-n-butyl phthalate; MBzP, Mono-benzyl phthalate; MCNP, Mono(carboxynonyl) phthalate; MCOP, Mono(carboxyoctyl) phthalate; MCPP, Mono-(3-carboxypropyl) phthalate; MECPP, Mono-2-ethyl-5-carboxypentyl phthalate; MEHHP, Mono-(2-ethyl-5-hydroxyhexyl) phthalate; MEHP, Mono-(2-ethyl)-hexyl phthalate; MEOHP, Mono-(2-ethyl-5-oxohexyl) phthalate; MEP, Mono-ethyl phthalate; MiBP, Mono-isobutyl phthalate; MP, Methyl paraben; PAH, polycyclic aromatic hydrocarbons; PP, Propyl paraben; 1-Naphthol, 1-hydroxynaphthalene; 1-OH-Phe, 1-hydroxyphenanthrene; 1-OHP, 1-hydroxypyrene; 2-Naphthol, 2-hydroxynaphthalene; 2-OH-Flu, 2-hydroxyfluorene; 2-OH-Phe, 2-hydroxyphenanthrene; 2,4-DCP, 24-dichlorophenol; 2,5-DCP, 25-dichlorophenol; 3-OH-Flu, 3-hydroxyfluorene; 3-OH-Phe, 3-hydroxyphenanthrene; 9-OH-Flu, 9-hydroxyfluorene.

Participants with the highest ERS_OS_ values (Q4: ≥26.05 IU/L), were on average older, more often male (83.4%), and were more likely to be Hispanic or Non-Hispanic Black, be a current or former smoker, and to have lower education attainment compared to participants in the lowest environmental risk for oxidative stress category (ERS_OS_ Q1: 16.95 IU/L) (**Table 3**). In addition, the highest ERS_OS_ group had lower HEI scores (lower adherence to the dietary guidelines),^30^ higher BMI, greater caloric and alcohol intakes and higher concentrations of GGT and cotinine. Finally, in the highest ERS_OS_ category (Q4) mortality cases were more frequent compared to the group with the lowest ERS_OS_ values (**Table 3**).

**Table 3:**
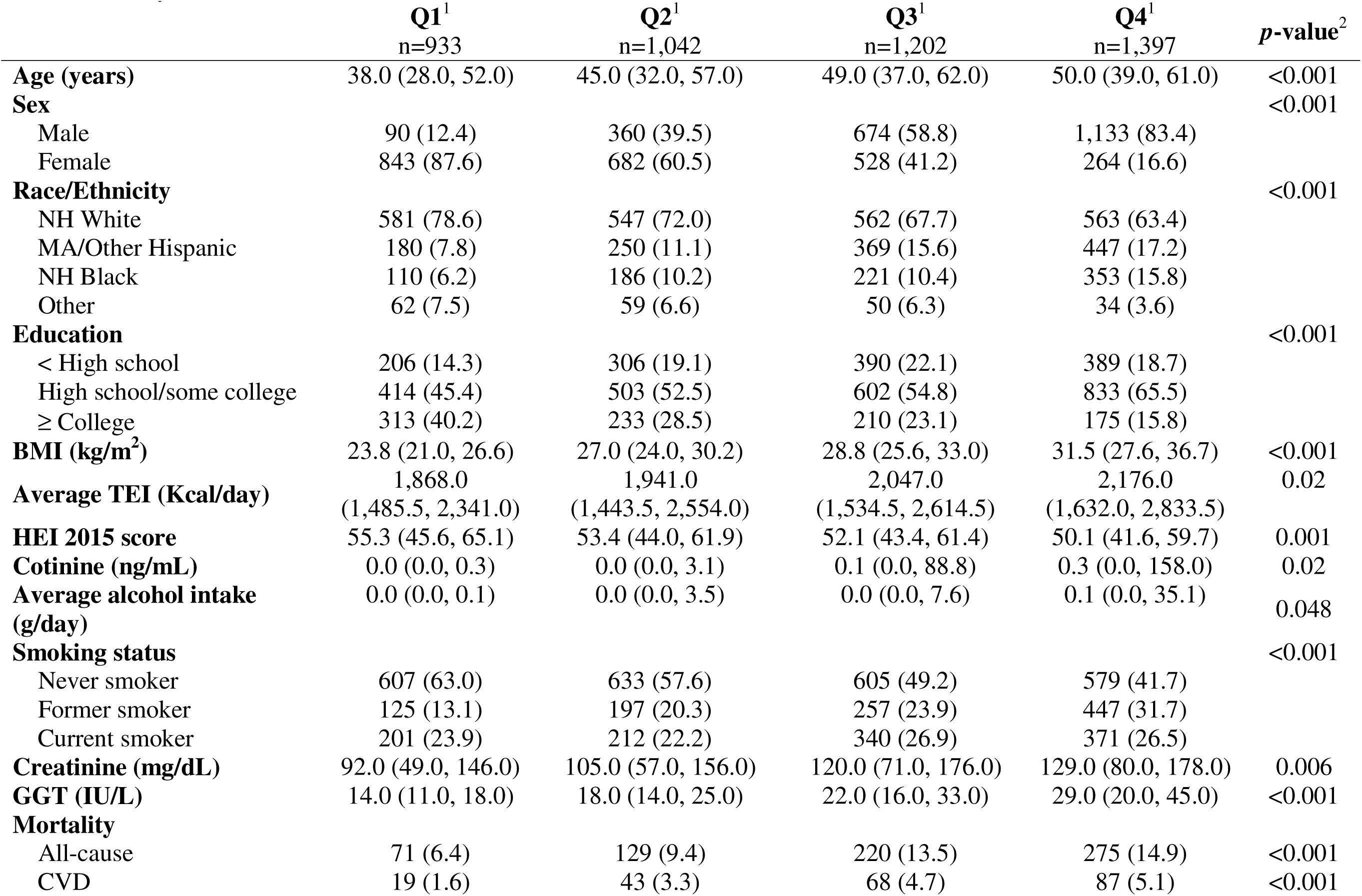
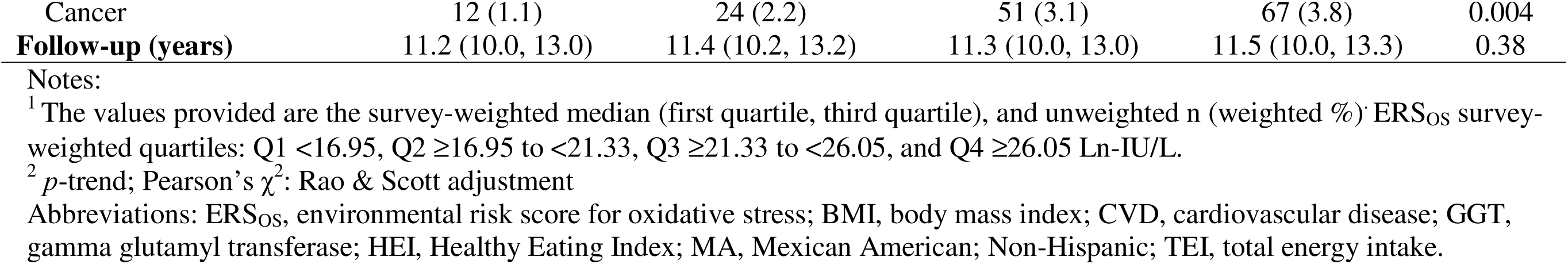
Population characteristics by ERS_OS_ survey-weighted quartiles in the National Health and Nutrition Examination Survey (NHANES) cycles 2005-2010 (n=4,574).

### Assessment of predictive performance of ERS_OS_

The median correlation between observed log-transformed GGT and predicted GGT (ERS_OS_) in the training sets was 0.46 (2.5^th^, 97.5^th^ percentiles: 0.44, 0.48). Slightly lower median correlation coefficient of 0.43 (2.5^th^, 97.5^th^ percentiles: 0.40, 0.48) was obtained in the testing sets. The MSE was marginally lower (median = 0.35 [2.5^th^, 97.5^th^ percentiles: 0.33, 0.38]) than the MSPE (median = 0.37 [2.5^th^, 97.5^th^ percentiles: 0.35, 0.39]) across 1,000 splits, reflecting a stable model with limited performance variance between training and testing sets.

#### Associations between ERS_OS_ and mortality

Covariate adjusted Cox proportional hazard models for all-cause, CVD, and cancer mortality were performed. A one-SD increase in the ERS_OS_ was associated with a 1.60-fold higher hazard of CVD mortality (empirical 2.5^th^, 97.5^th^ percentiles: 1.01, 2.57). Contrarily, ERS_OS_ quartiles were not associated with CVD mortality. No associations were observed for all-cause and cancer mortality when ERS_OS_ was modeled continuously or as quartiles (**Table 4**).

**Table 4:**
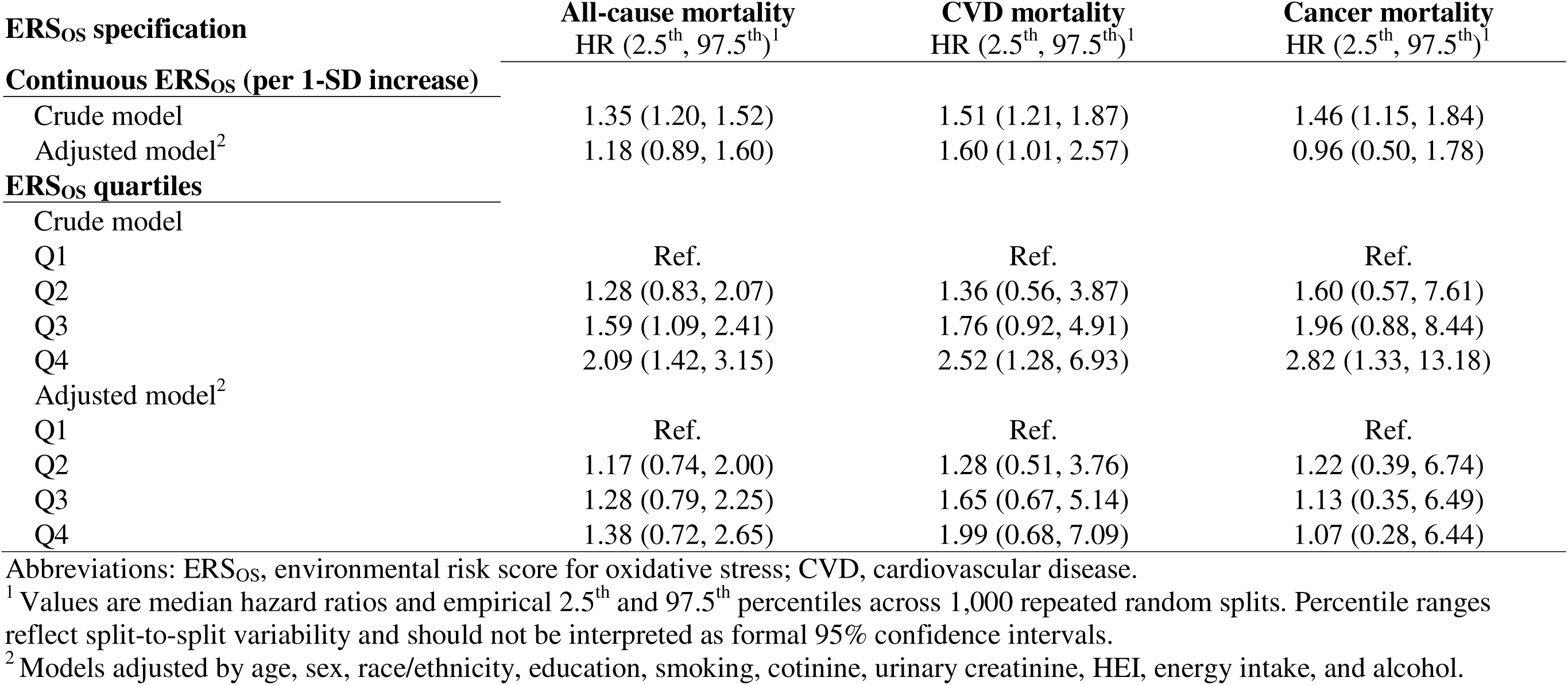
Proportional Hazards models between ERS_OS_ and mortality outcomes across 1000 testing sets.

#### Sensitivity analyses

Sensitivity analyses using a complete-case approach in the testing set yielded estimates of similar magnitude to our primary analysis. The positive association between ERS_OS_ and CVD mortality in the complete case analysis was slightly attenuated, with a one-SD increase in ERS_OS_ was associated with a 1.57-fold higher CVD mortality (median HR=1.57; empirical 2.5^th^, 97.5^th^ percentiles: 0.96, 2.90; **Table S5**).

## Discussion

To estimate the joint effect of multiple chemicals on oxidative stress, a common mechanism of toxicity, we constructed a biology-based ERS for oxidative stress, or ERS_OS_. The ERS_OS_ linked 30 chemicals across five toxicant classes including metals, PAHs, chlorophenol pesticides, phenols, and phthalates, measured in NHANES 2005-2010, using Qgcomp. We observed that higher ERS_OS_ values were associated with increased CVD mortality, suggesting that higher exposure to toxicants with high oxidative stress potential may increase the risk of CVD mortality. However, no associations were found for all-cause or cancer mortality.

Oxidative stress plays a central role in endothelial dysfunction, monocyte invasion, inflammation, atherosclerotic plaque formation, and in alterations in vasotone and vascular responses, all of which are mechanisms underlying CVD events.^38^ In contrast, cancer development often entails longer latency periods and involves multiple pathways beyond oxidative stress and chronic inflammation.^39^ Mechanisms such as genomic instability, epigenetic alterations, immune evasion and resistance to apoptosis interact with oxidative pathways but can also drive cancer development independently of oxidative stress.^39, 40^

The absence of an association with all-cause and cancer mortality across data splits may reflect etiologic heterogeneity relevant to carcinogenesis and other non-CVD deaths. Furthermore, a reduced number of cases may have limited statistical power to detect cancer mortality associations. For example, Zhang et al.^4^ examined a larger cohort of 8,378 adults from NHANES 2005-2016 and observed positive associations between a mixture of eight phthalates (MEP, MBP, MiBP, MBzP, MEHP, MEHHP, MEOHP, MECPP) and BPA and mortality outcomes. Using Qgcomp Cox proportional hazard models, they estimated that each tertile increase in this plasticizer mixture was associated with higher hazards of all-cause mortality (HR=1.35 [95% CI: 1.02, 1.78]), cancer mortality (HR = 1.79 [95% CI: 1.06, 3.03]), and CVD mortality (HR=1.83 [95% CI: 1.04, 3.22]).^4^ The association between the plasticizer mixture and CVD mortality reported by Zhang et al. was comparable to ours when ERS_OS_ was standardized (median HR=1.60 [empirical 2.5^th^, 97.5^th^ percentiles: 1.01, 2.57]). Moreover, although Zhang et al. used Qgcomp to model CVD mortality rather than GGT, MEOHP similarly obtained the largest negative weight.^4^

The ERS_OS_ creation using GGT as an outcome assumed the 30 chemical mixture components act, at least partially, through a shared common biological pathway, oxidative stress. Although oxidative stress is a pervasive mechanism underlying environmental chemical toxicity,^11^ toxicants can also exert effects through other pathways, such as inflammation, endocrine disruption, mitochondrial dysfunction, epigenetic changes, genomic alterations and mutations, among others.^2^ To address the complexity of mixture toxicity, this biology-informed index was motivated by the TEQ framework established for dioxin and dioxin-like chemicals. TEQ employs toxicity equivalency factors (TEFs), a normalization method in which the health risks of individual chemicals within a mixture are scaled against a common reference (i.e. the chemical with the greatest toxicity) to reflect differential toxic potency.^5^ TEFs are primarily determined based on each compound’s ability to bind and activate the aryl hydrocarbon receptor (AhR) and subsequent AhR-mediated toxic responses.^12^ In epidemiological studies, TEQs have been used evaluate the associations between dioxin and dioxin-like compounds among diverse health outcomes, such as reproductive and developmental endpoints,^41^ thyroid hormone levels,^42^ and mortality,^43^ by collapsing the measurements of these compounds into a single, biologically meaningful exposure. For example, a study in the European Prospective Investigation into Cancer and Nutrition (EPIC) cohort, comprising 451,390 adults across nine countries, observed a U-shaped non-linear associations between all-cause mortality, and dietary intakes of 17 dioxins and 12 dioxin-like polychlorinated biphenyls (DL-PCBs) expressed as TEQ.^43^ The use of TEQ in that study allowed for the characterization of the joint toxic burden of diverse dioxin and dioxin-like and the evaluation of their dose-response relationships across mortality outcomes.

Similar strategies have been applied to phthalate mixtures, combining exposure concentrations according to antiandrogenic^44^ or estrogenic activity.^45^ TEQ is calculated as the sum of the products of each compound’s TEF and its exposure concentration. The use of biology-driven exposure metrics could enhance the interpretability and translational relevance of mixture analyses in environmental epidemiology.

The interpretation of Qgcomp weights further illustrates the complexity of mixture effect analyses. In settings where directional homogeneity does not hold, Qgcomp weights reflect each chemical’s proportional contribution to the overall mixture effect in a given direction, with all weights in the same direction constrained to add to 1.0. Therefore, the magnitude of a chemical’s weight can only be compared with other chemicals in the same direction.^9^ In the conditional Qgcomp model, metabolites within the same chemical class often showed opposing associations with GGT, despite sharing common sources and being positive correlated. This heterogeneous pattern was observed among phenols and parabens, phthalates, and PAHs. Blood metals and chlorophenol pesticides were the only chemical classes with consistently positive Qgcomp weights. For example, among DEHP metabolites, MEHHP, and MECPP were positively associated with GGT, whereas MEOHP, and MEHP showed inverse associations (**Figure 2**). These findings contrast with the single-pollutant regressions, in which MEHHP and MECPP showed modest positive associations with GGT, while for the remaining DEHP metabolites (MEOHP, MEHP) no associations were observed. This discrepancy underscores how Qgcomp can yield effect estimates that diverge in direction and magnitude from single-exposure analyses, reflecting the joint modeling of correlated chemicals rather than isolated effects. Chemicals with modest individual associations may receive disproportionately large weights, highlighting the need for caution when interpreting Qgcomp weights as indicators of chemical-specific toxicity.

Comprehensive regulatory strategies that extend beyond single-chemical exposures and target groups of chemicals, such as the case of per- and polyfluoroalkyl substances (PFAS),^46^ could allow for more effective overall risk reduction as it would include those chemicals with individually small effects, but meaningful joint contributions. Nonetheless, the contradictory effect directions observed within chemical classes raise important questions regarding the use of mixture methods that rely on directional homogeneity assumptions, especially when evaluating complex chemical mixture comprising multiple toxicant classes. Another aspect to consider is the impact chemical concentrations have in the observed effects, as a chemical’s toxicity is a function of its exposure concentration and duration. The concentrations of the two DEHP metabolites positively associated with GGT (i.e. MEHHP and MECP) were up to 13 times higher to the concentrations of MEOHP and MEHP. However, the current approach, fails to account for chemical concentration. It is possible the inverse associations observed between GGT, MEOHP and MEHP could be attributed to their lower concentrations compared to metabolites derived for the same parent compound.

Several study limitations warrant consideration. First, due to availability of exposure data measured in the same participants in NHANES 2005-2010, we were able to include only five chemical classes. The presence of unmeasured chemicals may have resulted in residual confounding and biased weighting of mixture components when using Qgcomp. Future chemical mixture studies would benefit from incorporating a broader range of environmental exposures that better characterize environmental exposure profiles. Second, although GGT is commonly used as an oxidative stress measure in epidemiological studies, it remains a surrogate biomarker,^16, 17^ which could have introduced outcome misclassification. Plasma 8-iso-prostaglandin F2α (8-iso-PGF2α) is currently regarded as the gold standard for in vivo oxidative stress.^47^ Third, the reliance on a single GGT measurement may not have adequately reflected long-term oxidative stress. Fourth, the use of single-measurement exposure assessment prevented the evaluation of cumulative or time-varying exposures, which may be particularly relevant for mortality outcomes. Although repeated biomarker measurements could better capture chronic biological responses and dynamic exposure profiles, integrating longitudinal data to mixture analyses would introduce additional analytic complexity. Finally, although Qgcomp allows flexible a-priori specification of nonlinear terms or interactions, it remains a parametric approach as it assumes linear and additive exposure-response relationships.^9^ Alternative mixture methods, such as Bayesian kernel machine regression (BKMR), provide semiparametric frameworks capable of evaluating the independent or joint effect of the mixture components on an outcome while accounting for non-linear and non-additive relationships.^48^ However, although BKMR may offer a more flexible characterization of mixture effects, its current inability to incorporate survey weights limits its application to complex survey designs. Methodological advances that integrate mixture modeling with complex sampling structures, may improve the generalizability of environmental mixture analyses.

Despite these limitations, this study has notable strengths. We examined biomarkers from multiple chemical classes that are rarely studied jointly as part of a mixture within a nationally representative sample. Although the number of chemical classes was limited, few nationally representative studies have applied mixture methods across multi-class biomarkers. To achieve this, we used Qgcomp, a method useful in settings where prior knowledge of the direction and magnitude of individual chemical effects is limited, due to its flexibility regarding directionality homogeneity assumptions.^9^ Furthermore, unlike other chemical mixture studies using national data, the current analysis using survey-weighted Qgcomp incorporated sampling weights and clusters accounting for the multistage sampling design of NHANES,^9^ strengthening the validity and generalizability of the findings. Finally, we used the repeated random training/testing splits to construct ERS_OS_ and evaluate its association with mortality, providing more robust estimates. In this study we constructed a biology-based ERS by integrating the oxidative stress potential (as measured by their effect on GGT) of a multi-class chemical mixture in a representative US sample. The proposed approach grouped chemicals according to a shared biological mechanism, oxidative stress, rather than solely by structure or chemical class, while accounting for complex survey design features within a quantile g-computation framework. While this strategy assumes common pathways, biologically supported mixture analyses may offer an interpretable framework when mechanistic evidence supports common toxicity pathways. This integrated measure of oxidative stress (ERS_OS_) was associated with higher CVD mortality risk. However, the ERS_OS_ framework could be extended to alternative biological pathways by constructing risk scores based on other biomarkers relevant to chemical toxicity and applying them to downstream health outcomes beyond mortality. Finally, the use of Qgcomp with sampling weights and clusters may serve as an early analytic framework for future chemical mixture studies seeking to integrate chemical mixtures based on common biological pathways while accounting for complex sampling designs. Such biologically informed risk metrics may facilitate integration of mechanistic toxicology with population-level epidemiology.

## Supporting Information

Supplementary tables and figures: additional descriptive statistics, supporting visualizations, and sensitivity analyses (PDF).

## Supporting information

Supplementary tables and figures: additional descriptive statistics, supporting visualizations, and sensitivity analyses (PDF)

## Data Availability

Code produced in the present study is available upon reasonable request to the authors

## Acknowledgements

This study was supported by grants from the National Institute on Aging [R01-AG070897, K01-AG084821] and the National Institute of Environmental Health Sciences [P30-ES017885].

