## Supplementary tables and figures: additional descriptive statistics, supporting visualizations, and sensitivity analyses (PDF) for "Development of a Biology-Informed Chemical Mixture Index for Oxidative Stress and Mortality in NHANES 2005-2010: A Survey-Weighted Quantile G-Computation Approach"

**Supplementary Table S1:** Population characteristics of the excluded and included individuals from the National Health and Nutrition Examination Survey (NHANES) cycles 2005-2010 (n=31,034).

| | **Overall**[1] N = 31,034 | **Excluded**[1] N= 26,460 | **Included**[1] N = 4,574 | p-value[2] |
|---|---|---|---|---|
| **GGT (IU/L)** | 19.0 (13.0, 29.0) | 18.0 (13.0, 28.0) | 20.0 (14.0, 30.0) | <0.001 |
| **Age (years)** | 36 (17.0, 53.0) | 32 (14.0, 51.0) | 46 (33.0, 58.0) | <0.001 |
| **Female** | 15,633.0 (51.0) | 13,316.0 (51.0) | 2,317.0 (51.0) | 0.96 |
| **Race/Ethnicity** | | | | <0.001 |
| NH White | 12,463 (66.4) | 10,210 (65.4) | 2,253 (70.4) | |
| MA/Other Hispanic | 10,071 (14.8) | 8,825 (15.3) | 1,246 (12.9) | |
| NH Black | 6,878 (12.2) | 6,008 (12.6) | 870 (10.7) | |
| Other | 1,622 (6.5) | 1,417 (6.7) | 205 (6.0) | |
| **Education** | | | | 0.35 |
| < High school | 5,031 (19.1) | 3,742 (19.4) | 1,289 (18.4) | |
| High school/some college | 8,733 (54.6) | 6,384 (54.6) | 2,349 (54.4) | |
| < High school | 3,337 (26.3) | 2,407 (26.0) | 930 (27.1) | |
| **BMI (kg/m$^2$)** | 25.8 (21.4, 30.6) | 25.3 (20.7, 30.1) | 27.5 (24.0, 32.0) | <0.001 |
| **Average energy intake (Kcal/day)** | 1,913.0 (1,459.5, 2,474.5) | 1,891.0 (1,443.0, 2,446.5) | 1,998.0 (1,526.5, 2,576.5) | <0.001 |
| **HEI 2015 score** | 50.8 (42.4, 60.3) | 50.5 (42.1, 59.8) | 52.9 (43.6, 62.0) | <0.001 |
| **Cotinine (ng/mL)** | 0.1 (0.0, 2.4) | 0.1 (0.0, 1.6) | 0.1 (0.0, 33.2) | 0.09 |
| **Average alcohol intake (g/day)** | 0.0 (0.0, 0.2) | 0.0 (0.0, 0.0) | 0.0 (0.0, 9.4) | <0.001 |
| **Smoking status** | | | | 0.72 |
| Never smoker | 9,104 (53.3) | 6,682 (53.5) | 2,422 (52.8) | |
| Former smoker | 3,754 (22.3) | 2,728 (22.3) | 1,026 (22.4) | |
| Current smoker | 4,261 (24.4) | 3,138 (24.2) | 1,123 (24.9) | |
| **Creatinine (mg/dL)** | 109.0 (63.0, 165.0) | 109.0 (63.0, 165.0) | 111.0 (63.0, 165.0) | 0.82 |
| **Mortality outcomes** | | | | |
| All-cause | 3,014 (12.2) | 2,319 (12.6) | 695 (10.9) | 0.02 |
| CVD | 921 (3.8) | 704 (3.9) | 217 (3.6) | 0.47 |
| Cancer | 678 (3.0) | 524 (3.3) | 154 (2.5) | 0.02 |
| **Follow-up (years)** | 11.4 (10.1, 13.3) | 11.5 (10.0, 13.3) | 11.3 (10.1, 13.1) | 0.42 |

[1] The values provided are the survey-weighted median (first quartile, third quartile), and unweighted n (weighted %)

[2] Design-based Kruskal-Wallis test; Pearson's $\chi^2$: Rao & Scott adjustment**.**

Abbreviations: BMI, body mass index; CVD, cardiovascular disease; GGT, gamma glutamyl transferase; HEI, Healthy Eating Index; MA, Mexican American; NH, Non-Hispanic; TEI, total energy intake.

**Supplementary Table S2:** Population characteristics by mortality outcomes in the National Health and Nutrition Examination Survey (NHANES) cycles 2005-2010 (n=4,574)

| | **Non-cases** n=3,879 | **All-cause mortality** n=695[1] | $p^2$ | **CVD mortality** n=217[1] | $p^2$ | **Cancer mortality** n=154[1] | $p^2$ |
|---|---|---|---|---|---|---|---|
| **Age (years)** | 44.0 (32.0, 54.0) | 72.0 (60.0, 80.0) | <0.001 | 76.0 (65.0, 80.0) | <0.001 | 66.0 (54.0, 77.0) | <0.001 |
| **Sex** | | | 0.52 | | 0.68 | | 0.07 |
| Male | 1,855 (48.4) | 402 (50.2) | | 126 (50.2) | | 98 (59.1) | |
| Female | 2,024 (51.6) | 293 (49.8) | | 91 (49.8) | | 56 (40.9) | |
| **Race/Ethnicity** | | | <0.001 | | 0.006 | | 0.03 |
| NH White | 1,794 (69.1) | 459 (81.2) | | 150 (82.9) | | 90 (74.8) | |
| MA/Other Hispanic | 1,137 (13.7) | 109 (6.9) | | 29 (6.3) | | 27 (6.7) | |
| NH Black | 758 (10.8) | 112 (9.3) | | 35 (8.4) | | 35 (16.3) | |
| Other | 190 (6.4) | 15 (2.6) | | 3 (2.5) | | 2 (2.2) | |
| **Education** | | | <0.001 | | <0.001 | | 0.004 |
| < High school | 1,025 (16.9) | 266 (31.5) | | 81 (33.0) | | 59 (32.6) | |
| High school/some college | 2,017 (54.7) | 335 (53.0) | | 104 (51.0) | | 76 (51.3) | |
| ≥ College | 837 (28.3) | 94 (15.5) | | 32 (16.0) | | 19 (16.1) | |
| **BMI (kg/m²)** | 27.5 (23.9, 32.0) | 27.9 (24.4, 32.1) | 0.33 | 28.0 (24.1, 31.5) | 0.69 | 28.1 (24.6, 32.9) | 0.21 |
| **Avg TEI (Kcal/day)** | 2,025.0 (1,543.5, 2,612.0) | 1,733.0 (1,355.0, 2,197.0) | <0.001 | 1,610.5 (1,246.0, 2,003.0) | <0.001 | 1,805.0 (1,438.5, 2,264.0) | 0.007 |
| **HEI 2015 score** | 52.8 (43.5, 62.0) | 53.5 (45.2, 62.6) | 0.13 | 52.9 (43.2, 61.3) | 0.72 | 55.3 (45.8, 62.6) | 0.32 |
| **Cotinine (ng/mL)** | 0.1 (0.0, 27.3) | 0.1 (0.0, 67.2) | 0.86 | 0.0 (0.0, 1.0) | 0.28 | 0.1 (0.0, 86.4) | 0.80 |
| **Avg alcohol intake (g/day)** | 0.0 (0.0, 10.1) | 0.0 (0.0, 0.5) | 0.003 | 0.0 (0.0, 0.0) | 0.002 | 0.0 (0.0, 0.0) | 0.02 |
| **Smoking status** | | | <0.001 | | <0.001 | | 0.02 |
| Never smoker | 2,162 (54.6) | 262 (38.5) | | 81 (36.9) | | 53 (38.9) | |
| Former smoker | 873 (22.1) | 153 (23.3) | | 44 (21.3) | | 39 (23.2) | |
| Current smoker | 844 (23.2) | 280 (38.2) | | 92 (41.8) | | 62 (37.9) | |
| **Urinary creatinine (mg/dL)** | 114.0 (64.0, 168.0) | 91.0 (53.0, 130.0) | <0.001 | 86.0 (54.0, 127.0) | <0.001 | 101.0 (52.0, 152.0) | 0.03 |
| **GGT (IU/L)** | 20.0 (14.0, 30.0) | 21.0 (15.0, 35.0) | <0.001 | 20.0 (14.0, 33.0) | 0.59 | 20.0 (17.0, 32.0) | 0.06 |
| **Follow-up (years)** | 11.6 (10.3, 13.3) | 6.7 (3.7, 9.6) | <0.001 | 6.4 (3.4, 9.2) | <0.001 | 7.0 (4.3, 9.8) | <0.001 |

[1] Median (Q1, Q3); n (unweighted) (weighted %)

[2] Design-based Kruskal-Wallis test; Pearson's $\chi^2$: Rao & Scott adjustment

Abbreviations: Avg, average; GGT, gamma glutamyl transferase; HEI, Healthy Eating Index; MA, Mexican American; NH, Non-Hispanic; TEI, total energy intake.

**Supplementary Table S3:** Distributions of chemical biomarker concentrations by mortality outcomes in National Health and Nutrition Examination Survey (NHANES) cycles 2005-2010 (n=4,574).

| | **Non-cases**[1] n=3,879 | **All-cause mortality**[1] n=695 | *p*[2] | **CVD mortality**[1] n=217 | *p*[2] | **Cancer mortality**[1] n=154 | *p*[2] |
|---|---|---|---|---|---|---|---|
| **Blood metals** | | | | | | | |
| Lead (ug/dL) | 1.2 (0.8, 1.9) | 1.9 (1.3, 2.8) | <0.001 | 2.1 (1.5, 3.2) | <0.001 | 1.8 (1.3, 2.6) | <0.001 |
| Cadmium (ug/L) | 0.3 (0.2, 0.6) | 0.5 (0.3, 0.8) | <0.001 | 0.5 (0.3, 0.8) | <0.001 | 0.5 (0.3, 0.9) | <0.001 |
| Mercury total (ug/L) | 1.0 (0.5, 2.1) | 0.9 (0.5, 1.6) | <0.001 | 1.0 (0.5, 1.5) | 0.03 | 0.9 (0.4, 1.6) | 0.06 |
| **Chlorophenol pesticides**, ug/L | | | | | | | |
| 2,4-DCP | 0.8 (0.3, 1.7) | 0.7 (0.3, 1.5) | 0.47 | 0.7 (0.4, 1.5) | 0.87 | 0.6 (0.3, 1.4) | 0.08 |
| 2,5-DCP | 6.0 (1.9, 21.4) | 6.4 (1.9, 22.3) | 0.85 | 7.0 (2.0, 25.0) | 0.74 | 7.6 (1.8, 23.4) | 0.71 |
| **PAH**, ng/L | | | | | | | |
| 1-Naphthol | 2,004.0 (840.3, 7,335.3) | 3,010.0 (969.0, 10,320.0) | 0.002 | 2,026.2 (834.4, 8,573.9) | 0.71 | 3,285.0 (969.0, 10,815.0) | 0.04 |
| 2-Naphthol | 3,708.0 (1,656.6, 8,775.0) | 3,254.0 (1,337.1, 8,414.0) | 0.08 | 2,633.4 (1,369.7, 8,861.3) | 0.03 | 4,736.0 (1,195.0, 9,501.0) | 0.91 |
| 2-OH-Flu | 253.8 (123.0, 653.2) | 204.0 (109.0, 659.0) | 0.11 | 189.0 (105.7, 491.2) | 0.02 | 204.0 (121.0, 612.7) | 0.42 |
| 3-OH-Flu | 90.0 (42.0, 291.3) | 67.0 (34.6, 279.7) | 0.01 | 53.9 (31.0, 213.4) | 0.001 | 68.1 (35.7, 263.0) | 0.25 |
| 9-OH-Flu | 308.0 (151.9, 626.0) | 298.0 (155.0, 627.0) | 0.99 | 285.5 (138.0, 623.7) | 0.67 | 274.0 (136.0, 577.0) | 0.46 |
| 1-OH-Phe | 146.1 (78.9, 265.0) | 128.3 (72.1, 237.8) | 0.048 | 120.9 (66.0, 237.8) | 0.02 | 153.0 (63.1, 252.1) | 0.48 |
| 2-OH-Phe | 67.4 (36.8, 126.9) | 60.2 (31.2, 110.0) | 0.04 | 58.5 (29.0, 107.4) | 0.03 | 55.0 (30.0, 117.0) | 0.28 |
| 3-OH-Phe | 86.0 (43.9, 171.4) | 79.1 (38.9, 156.0) | 0.16 | 70.0 (35.4, 148.9) | 0.07 | 79.1 (38.6, 175.9) | 0.49 |
| 1-OHP | 109.0 (52.0, 222.0) | 68.0 (34.3, 163.3) | <0.001 | 60.4 (33.2, 135.1) | <0.001 | 73.0 (33.3, 191.0) | 0.009 |
| **Phenols and parabens**, ng/mL | | | | | | | |
| BPA | 1.9 (0.9, 3.7) | 1.8 (0.9, 3.5) | 0.15 | 1.7 (0.7, 2.9) | 0.07 | 1.9 (0.9, 3.9) | 0.81 |
| BP-3 | 16.3 (4.5, 80.5) | 6.9 (1.6, 39.5) | <0.001 | 6.6 (1.5, 31.2) | <0.001 | 6.4 (1.6, 39.5) | 0.005 |
| Triclosan | 13.8 (3.3, 77.4) | 7.8 (1.6, 29.2) | <0.001 | 11.5 (1.6, 33.6) | 0.03 | 7.8 (2.4, 32.0) | 0.03 |
| Methyl paraben | 64.2 (16.2, 220.0) | 48.4 (11.3, 208.0) | 0.046 | 63.6 (15.6, 167.0) | 0.62 | 32.1 (7.5, 176.0) | 0.03 |
| Propyl paraben | 9.3 (1.3, 50.3) | 4.8 (0.6, 41.0) | <0.001 | 5.3 (0.8, 54.4) | 0.23 | 4.3 (0.4, 29.4) | 0.02 |
| **Phthalates**, ng/mL | | | | | | | |
| MEHHP | 16.8 (7.6, 38.6) | 14.6 (6.6, 31.7) | 0.01 | 15.7 (7.1, 29.2) | 0.27 | 13.2 (6.0, 37.9) | 0.12 |
| MEOHP | 10.1 (4.7, 22.4) | 9.5 (4.0, 19.1) | 0.02 | 10.2 (4.3, 18.3) | 0.42 | 9.2 (3.8, 23.4) | 0.12 |
| MEHP | 1.9 (0.9, 5.0) | 1.2 (0.8, 2.9) | <0.001 | 1.3 (0.9, 2.7) | 0.003 | 1.4 (0.8, 2.9) | <0.001 |
| MECPP | 25.2 (12.1, 54.5) | 23.5 (11.0, 49.9) | 0.09 | 26.0 (11.6, 49.1) | 0.88 | 20.0 (9.3, 52.7) | 0.04 |
| MBP | 16.8 (8.3, 32.6) | 16.3 (8.1, 31.0) | 0.79 | 16.2 (7.7, 31.0) | 0.78 | 17.6 (8.9, 29.2) | 0.96 |
| MiBP | 6.9 (3.2, 13.4) | 5.2 (2.3, 10.3) | <0.001 | 5.0 (2.2, 10.0) | 0.001 | 5.1 (2.4, 8.6) | 0.008 |
| MEP | 81.3 (30.1, 239.9) | 76.2 (29.2, 265.1) | 0.80 | 99.5 (35.9, 258.5) | 0.16 | 70.4 (29.4, 317.8) | 0.37 |

| | | | | | | | |
|---|---|---|---|---|---|---|---|
| MBzP | 6.7 (2.9, 15.0) | 6.0 (2.5, 12.9) | 0.04 | 5.0 (2.3, 11.7) | 0.09 | 6.8 (3.2, 15.2) | 0.78 |
| MCPP | 2.4 (1.1, 4.9) | 2.0 (0.9, 4.3) | 0.01 | 2.2 (0.9, 4.9) | 0.28 | 1.9 (0.9, 3.8) | 0.01 |
| MCNP | 2.5 (1.3, 5.1) | 2.2 (1.1, 4.4) | 0.08 | 2.4 (1.0, 5.5) | 0.33 | 1.7 (1.0, 4.0) | 0.006 |
| MCOP | 6.8 (3.1, 17.8) | 5.1 (2.5, 11.6) | <0.001 | 5.8 (2.4, 15.3) | 0.07 | 5.0 (2.6, 11.0) | 0.02 |

[1] Median (Q1, Q3)
[2] Design-based Kruskal-Wallis test
Notes:
The values provided are the survey-weighted median (first quartile, third quartile)
Metal biomarkers were measured in blood, whereas chlorophenol pesticides, PAH, phenols and parabens, and phthalates were measured in blood.
Abbreviations: BPA, Urinary Bisphenol A; BP-3, Urinary Benzophenone-3; CVD, cardiovascular disease; DEHP, Di(2-ethylhexyl) phthalate; MBP, Mono-n-butyl phthalate; MBzP, Mono-benzyl phthalate; MCNP, Mono(carboxynonyl) phthalate; MCOP, Mono(carboxyoctyl) phthalate; MCPP, Mono-(3-carboxypropyl) phthalate; MECPP, Mono-2-ethyl-5-carboxypentyl phthalate; MEHHP, Mono-(2-ethyl-5-hydroxyhexyl) phthalate; MEHP, Mono-(2-ethyl)-hexyl phthalate; MEOHP, Mono-(2-ethyl-5-oxohexyl) phthalate; MEP, Mono-ethyl phthalate; MiBP, Mono-isobutyl phthalate; MP, Methyl paraben; PAH, polycyclic aromatic hydrocarbons; PP, Propyl paraben; 1-Naphthol, 1-hydroxynaphthalene; 1-OH-Phe, 1-hydroxyphenanthrene; 1-OHP, 1-hydroxypyrene; 2-Naphthol, 2-hydroxynaphthalene; 2-OH-Flu, 2-hydroxyfluorene; 2-OH-Phe, 2-hydroxyphenanthrene; 2,4-DCP, 24-dichlorophenol; 2,5-DCP, 25-dichlorophenol; 3-OH-Flu, 3-hydroxyfluorene; 3-OH-Phe, 3-hydroxyphenanthrene; 9-OH-Flu, 9-hydroxyfluorene.

**Supplementary Figure S1:** Heatmap of unweighted Spearman correlation coefficients between chemical biomarkers in the National Health and Nutrition Examination Survey (NHANES) waves 2005-2010 in the full sample (n=4,574).

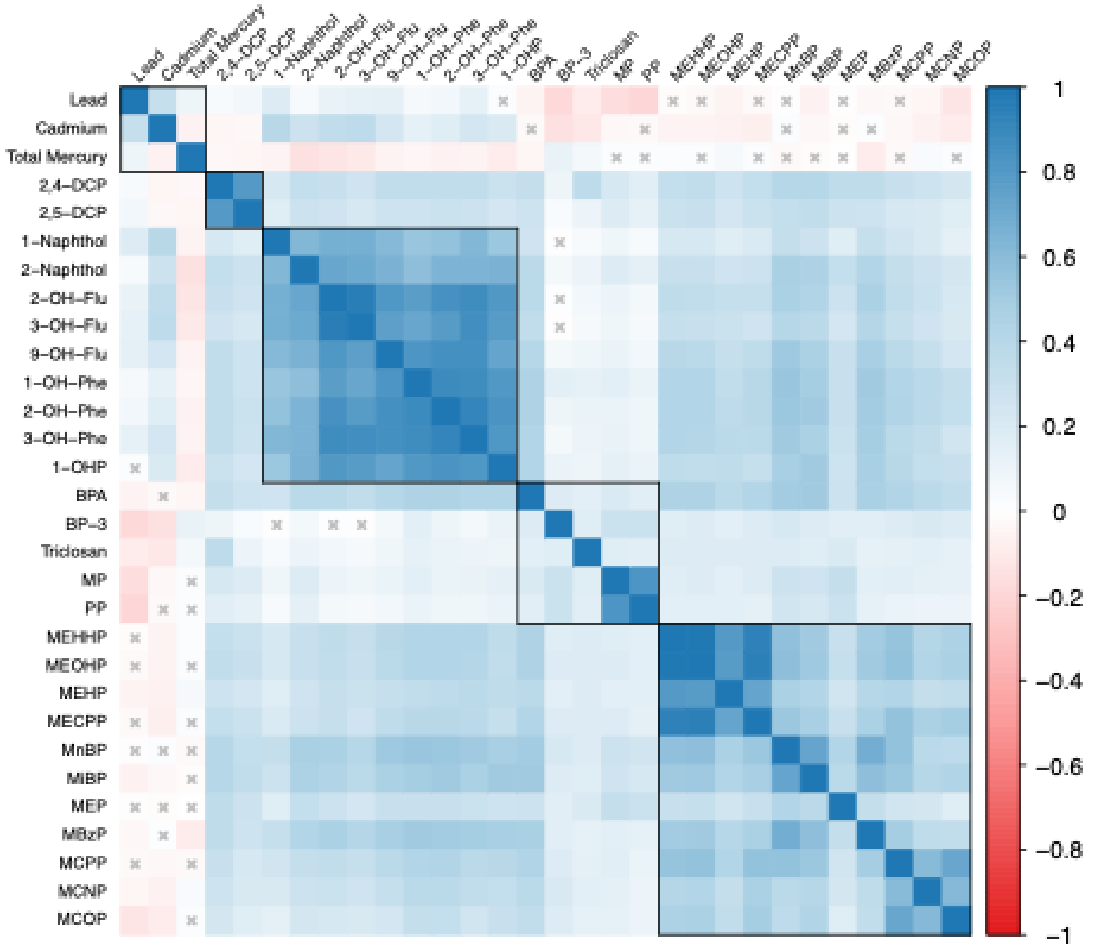


Notes: The shades of red and blue in the lower triangle indicate the strength of positive and negative correlations, respectively. Correlations with *p*-values >0.05 are marked with an "X". Blacked boxes outline the biomarkers by chemical class (blood metals, urinary chlorophenol pesticides, polycyclic aromatic hydrocarbons (PAH), phenols and parabens, and phthalates). Abbreviations: BPA, Urinary Bisphenol A; BP-3, Urinary Benzophenone-3; DEHP, Di(2-ethylhexyl) phthalate; MBP, Mono-n-butyl phthalate; MBzP, Mono-benzyl phthalate; MCNP, Mono(carboxynonyl) phthalate; MCOP, Mono(carboxyoctyl) phthalate; MCPP, Mono-(3-carboxypropyl) phthalate; MECPP, Mono-2-ethyl-5-carboxypentyl phthalate; MEHHP, Mono-(2-ethyl-5-hydroxyhexyl) phthalate; MEHP, Mono-(2-ethyl)-hexyl phthalate; MEOHP, Mono-(2-ethyl-5-oxohexyl) phthalate; MEP, Mono-ethyl phthalate; MiBP, Mono-isobutyl phthalate; MP, Methyl paraben; PP, Propyl paraben; 1-Naphthol, 1-hydroxynaphthalene; 1-OH-Phe, 1-hydroxyphenanthrene; 1-OHP, 1-hydroxypyrene; 2-Naphthol, 2-hydroxynaphthalene; 2-OH-Flu, 2-hydroxyfluorene; 2-OH-Phe, 2-hydroxyphenanthrene; 2,4-DCP, 24-dichlorophenol; 2,5-DCP, 25-dichlorophenol; 3-OH-Flu, 3-hydroxyfluorene; 3-OH-Phe, 3-hydroxyphenanthrene; 9-OH-Flu, 9-hydroxyfluorene.

**Supplementary Table S4:** Single-chemical survey-weighted regressions between chemical mixture components quartiles and log-transformed GGT in the training set (n=2,287).

| | β | 95% CI | *p* |
|---|---|---|---|
| **Metals (blood)** | | | |
| Lead | 0.055 | 0.017, 0.092 | **0.007** |
| Cadmium | 0.018 | -0.017, 0.054 | 0.32 |
| Mercury total | 0.020 | -0.014, 0.053 | 0.26 |
| **Chlorophenol pesticides (urine)** | | | |
| 2,4-DCP | -0.015 | -0.045, 0.014 | 0.31 |
| 2,5-DCP | -0.008 | -0.035, 0.020 | 0.58 |
| **PAH (urine)** | | | |
| 1-Naphthol | -0.006 | -0.041, 0.029 | 0.74 |
| 2-Naphthol | 0.026 | -0.008, 0.06 | 0.15 |
| 2-OH-Flu | -0.008 | -0.049, 0.033 | 0.70 |
| 3-OH-Flu | -0.046 | -0.091, -0.001 | 0.05 |
| 9-OH-Flu | 0.032 | -0.008, 0.072 | 0.13 |
| 1-OH-Phe | 0.002 | -0.038, 0.042 | 0.93 |
| 2-OH-Phe | -0.002 | -0.035, 0.031 | 0.90 |
| 3-OH-Phe | -0.034 | -0.074, 0.006 | 0.11 |
| 1-OHP | 0.002 | -0.034, 0.038 | 0.93 |
| **Phenols and parabens (urine)** | | | |
| BPA | -0.008 | -0.038, 0.021 | 0.59 |
| BP-3 | 0.003 | -0.027, 0.034 | 0.84 |
| Triclosan | 0.002 | -0.023, 0.027 | 0.90 |
| Methyl paraben | 0.001 | -0.032, 0.035 | 0.94 |
| Propyl paraben | -0.008 | -0.041, 0.024 | 0.63 |
| **Phthalates (urine)** | | | |
| MEHHP | 0.021 | -0.002, 0.044 | 0.09 |
| MEOHP | -0.001 | -0.022, 0.021 | 0.94 |
| MEHP | 0.002 | -0.027, 0.032 | 0.89 |
| MECPP | 0.032 | 0.008, 0.055 | **0.01** |
| MBP | 0.016 | -0.024, 0.057 | 0.44 |
| MiBP | 0.030 | -0.002, 0.061 | 0.07 |
| MEP | 0.008 | -0.020, 0.035 | 0.58 |
| MBzP | 0.023 | -0.008, 0.054 | 0.16 |
| MCPP | -0.004 | -0.034, 0.027 | 0.81 |
| MCNP | -0.030 | -0.067, 0.007 | 0.12 |
| MCOP | -0.020 | -0.060, 0.019 | 0.33 |

Abbreviations: BPA, Urinary Bisphenol A; BP-3, Urinary Benzophenone-3; DEHP, Di(2-ethylhexyl) phthalate; MBP, Mono-n-butyl phthalate; MBzP, Mono-benzyl phthalate; MCNP, Mono(carboxynonyl) phthalate; MCOP, Mono(carboxyoctyl) phthalate; MCPP, Mono-(3-carboxypropyl) phthalate; MECPP, Mono-2-ethyl-5-carboxypentyl phthalate; MEHHP, Mono-(2-ethyl-5-hydroxyhexyl) phthalate; MEHP, Mono-(2-ethyl)-hexyl phthalate; MEOHP, Mono-(2-ethyl-5-oxohexyl) phthalate; MEP, Mono-ethyl phthalate; MiBP, Mono-isobutyl phthalate;

MP, Methyl paraben; PAH, polycyclic aromatic hydrocarbons; PP, Propyl paraben; 1-Naphthol, 1-hydroxynaphthalene; 1-OH-Phe, 1-hydroxyphenanthrene; 1-OHP, 1-hydroxypyrene; 2-Naphthol, 2-hydroxynaphthalene; 2-OH-Flu, 2-hydroxyfluorene; 2-OH-Phe, 2-hydroxyphenanthrene; 2,4-DCP, 24-dichlorophenol; 2,5-DCP, 25-dichlorophenol; 3-OH-Flu, 3-hydroxyfluorene; 3-OH-Phe, 3-hydroxyphenanthrene; 9-OH-Flu, 9-hydroxyfluorene.

**Supplementary Table S5:** Proportional Hazards models between $ERS_{OS}$ and mortality outcomes in complete cases across 1000 testing sets.

| **$ERS_{OS}$ specification** | **All-cause mortality** HR ($2.5^{th}$, $97.5^{th}$)[1] | **CVD mortality** HR ($2.5^{th}$, $97.5^{th}$)[1] | **Cancer mortality** HR ($2.5^{th}$, $97.5^{th}$)[1] |
|---|---|---|---|
| **Continuous $ERS_{OS}$ (per 1-SD increase)** | | | |
| Crude model | 1.34 (1.19, 1.53) | 1.46 (1.17, 1.80) | 1.52 (1.16, 2.00) |
| Adjusted model[2] | 1.24 (0.92, 1.76) | 1.57 (0.96, 2.90) | 1.08 (0.55, 2.09) |
| **$ERS_{OS}$ quartiles** | | | |
| Crude model | | | |
| Q1 | Ref. | Ref. | Ref. |
| Q2 | 1.21 (0.81, 1.92) | 1.44 (0.62, 3.97) | 1.46 (0.53, 7.56) |
| Q3 | 1.44 (1.01, 2.31) | 1.78 (0.84, 4.66) | 1.81 (0.73, 8.30) |
| Q4 | 1.95 (1.38, 3.09) | 2.48 (1.26, 6.32) | 2.95 (1.33, 12.41) |
| Adjusted model[2] | | | |
| Q1 | Ref. | Ref. | Ref. |
| Q2 | 1.10 (0.70, 1.84) | 1.41 (0.52, 4.18) | 1.15 (0.36, 6.37) |
| Q3 | 1.19 (0.73, 2.12) | 1.67 (0.62, 5.25) | 1.08 (0.30, 5.55) |
| Q4 | 1.38 (0.75, 2.62) | 2.05 (0.59, 7.93) | 1.20 (0.28, 6.95) |

[1] Values are median hazard ratios and empirical $2.5^{th}$ and $97.5^{th}$ percentiles across 1,000 repeated random splits. Percentile ranges reflect split-to-split variability and should not be interpreted as formal 95% confidence intervals.
[2] Models adjusted by age, sex, race/ethnicity, education, smoking, cotinine, urinary creatinine, HEI, energy intake, and alcohol.
Abbreviations: $ERS_{OS}$, environmental risk score for oxidative stress; CVD, cardiovascular disease.
